# The necessary cooperation between governments and public in the fight against COVID-19: why non-pharmaceutical interventions may be ineffective

**DOI:** 10.1101/2020.08.17.20176347

**Authors:** Christielly Mendonça Borges, Marco Túlio Pacheco Coelho, José Alexandre Felizola Diniz-Filho, Thiago Fernando Rangel

## Abstract

The coronavirus disease (COVID-19) outbreak is the biggest public health challenge in the last 100 years. No successful pharmaceutical treatment is yet available, thus effective public health interventions to contain COVID-19 include social distancing, isolation and quarantine measures, however the efficiency of these containment measures varied among countries and even within states in the same country. Despite Brazil being deeply affected by coronavirus, the federal government never proposed a coordinated action to control COVID-19 and Brazilian states, which are autonomous, each imposed different containment measures. The state of Goiás declared strict social distancing measures in March 13, but gradually relaxed many of its first measures due specially to public pressure. Here we use a Susceptible-Infected-Recovered (SIR) model combined with Bayesian inference and a time-dependent spreading rate to assess how past state-level interventions affected the spread of COVID-19 in Goiás. The interventions succeeded in decreasing the transmission rate in the state, however, after the third intervention the rate remained positive and exponential. Thus, other stricter interventions were made necessary to avoid the growth of new cases and a collapse in the health system. Governmental interventions need to be taken seriously by the population in order for them have the proposed outcome. Our results reflect the population’s disregard with the measures imposed and the need for cooperation between governments and its citizens in the fight against COVID-19.

## Introduction

The coronavirus disease 2019 (COVID-19) outbreak caught the world by surprise, as in three months it went from a public health emergency of international concern to a global pandemic *(1)*. This is the first pandemic caused by a coronavirus, since the severe acute respiratory syndrome coronavirus 2 (SARS-COV-2) is highly transmissible and causes a pathogenic viral infection *(2)*. Human to human spread occurs mainly through respiratory droplets and contact routes *(2)*, but the virus can remain infections in aerosols and surfaces up to days *(3)*. While therapies and vaccines are still not available, preventions of disease spread and mitigation of the pandemic relies in non-pharmaceutical interventions (NPI) such as social distancing, isolation, face covering and quarantine measures *(4, 5)*.

Despite being recommended by the World Health Organization (WHO), the adoption of NPIs varied greatly between countries. The virus emerged first in China, where strict social distancing rules were enforced early and only three months later the spreading was contained *(6, 7)*. Timing of intervention during the outbreak can explain discrepancies in the number of deaths in European countries, where Italy had 525 deaths per million population in contrast to Germany’s 95 deaths per million population in the same month *(8)*. As of June, the three countries with the highest number of COVID-19 related deaths are the US, Brazil and the UK *(9)*, all countries where heads of state and government openly spoke against social distancing measures *(10–12)*.

The first confirmed COVID-19 case in Brazil was registered on February 25, in the city of São Paulo *(13)*. By March, it had already reached all 26 states and the Federal District *(14)*. By June 25 Brazil reported over one million confirmed cases and a total of 58,314 deaths *(9)*. The states with the higher and lower confirmed cases were São Paulo and Mato Grosso do Sul, with 275,145 and 7,965 cases, respectively *(9)*. States are autonomous under the Brazilian constitution *(15)*, nonetheless, there was no coordinated action to control COVID-19 by the federal government.

Most state governors enforced restrictive contact measures in mid-March, when the virus began spreading. However, the president of the Federal Government has been vocally against state-level social distancing policies, citing frequently his fear of an economic collapse *(16)*. In fact, president Bolsonaro passed a provisional measure in April which entrusted to the Union prerogatives concerning isolation, quarantine and the interdiction of locomotion, public services and essential activities during the pandemic *(17)*. This measure was quickly overruled by the Supreme Federal Court of Justice, instating the Union could legislate on the subject but must always safeguard the autonomy of states and municipalities *(17)*. This political confrontation deepened an already existing rift in the population, with Bolsonaro’s supporters positioning themselves against states and municipalities’ COVID-19 containment measures.

Brazil is a continental sized country with substantial regional socioeconomic inequalities, all factors that further reduce support for social distancing measures *(14)*, and results in different containment measures in different states. In April, while cities such as Manaus, Fortaleza, Brasília, Rio de Janeiro and São Paulo faced an exponential growth of COVID-19 cases *(18)*, southern states Santa Catarina and Rio Grande do Sul started to reopen their economies without many registered cases (Pellegrini, 2020; Silva et al., 2020 [in press]). The state of Goiás was amongst the leaders of social isolation, registering in March over 60% of reduced mobility monitored via geolocation in smartphones *(21)*. That percentage eventually dropped and, more recently, Goiás recorded only 37% of reduced mobility, one of the worst rates of isolation in the country *(21)*. As a consequence, cases and deaths started to increase fast.

The first confirmed COVID-19 case in Goiás was registered on March 12. By March 13, the state government issued a decree declaring public health emergency and instituted strict social distancing measures. However, due to public and economic pressure, the government gradually relaxed many of its first measures *(22)*. Thus, in this paper we seek to detect if change points in the effective growth rate of COVID-19 correlates with governmental interventions made in Goiás. We use a Susceptible-Infected-Recovered (SIR) model combined with Bayesian inference and a time-dependent spreading rate *(23)* to assess the early transmission dynamics and evaluate the effectiveness of the state-level interventions. Short-term forecasts such as this are key to estimate medical requirements and capacities, and here we use it to assess how past mitigations affected the spread of COVID-19 in the state.

## Methods

We reproduced the framework stablished by Dehning *et. al*. (2020, https://github.com/Priesemann-Group/covid19_inference). They combined SIR models with Bayesian parameter inference with Markov Chain Monte Carlo (MCMC) sampling and augmented the model by a time-dependent spreading rate, which is implemented via potential change points that characterize governmental interventions *(23)*. We adjusted the model, initially created for Germany, for the state of Goiás, by choosing the three main state-level interventions and parameters accordingly.

SIR models have been used broadly to model epidemic spreads *(24, 25)*, and recently gained strength in efforts to model the spread of COVID-19 worldwide *(23, 26, 27)*. SIR models specify the rates that population recover and become infected by a disease. Bayesian inference with MCMC sampling assimilates prior knowledge available and accounts for data uncertainties into forecasts. An integration between Bayesian inference and SIR models provide a better assessment of more complex and realistic models *(28)*. Here we ran (1) an SIR model for the initial onset period with stationary spreading rate (simple SIR model) and (2) a time-dependent SIR model with weekend correction (full SIR model) *(23)*.

### Goiás characterization and data

The state of Goiás is located in the mid-west region of Brazil and has a population of approximately 7 million people *(29)*. The Federal District, along with the country’s capital Brasília, is geographically embedded within the State of Goiás, but due to administrative differences and independence of public health policies, here we analyze only data for Goiás. Daily number of COVID-19 confirmed cases in Goiás came from the Goiás State Health Department (SES-GO; acronym in Portuguese). SES-GO systematically monitors suspected cases throughout the state and provides daily updates of confirmed cases *(30)*. We used data until May 22.

### Governmental interventions

As of May 22, Goiás had amounted 14 decrees regarding the coronavirus pandemic. Most of these decrees are relaxations of the first decree, such as reopening churches and temples. To implement and maintain the model simple, we chose the three main decrees capable of influencing public behavior (Fig. 1).

**Figure 1.**
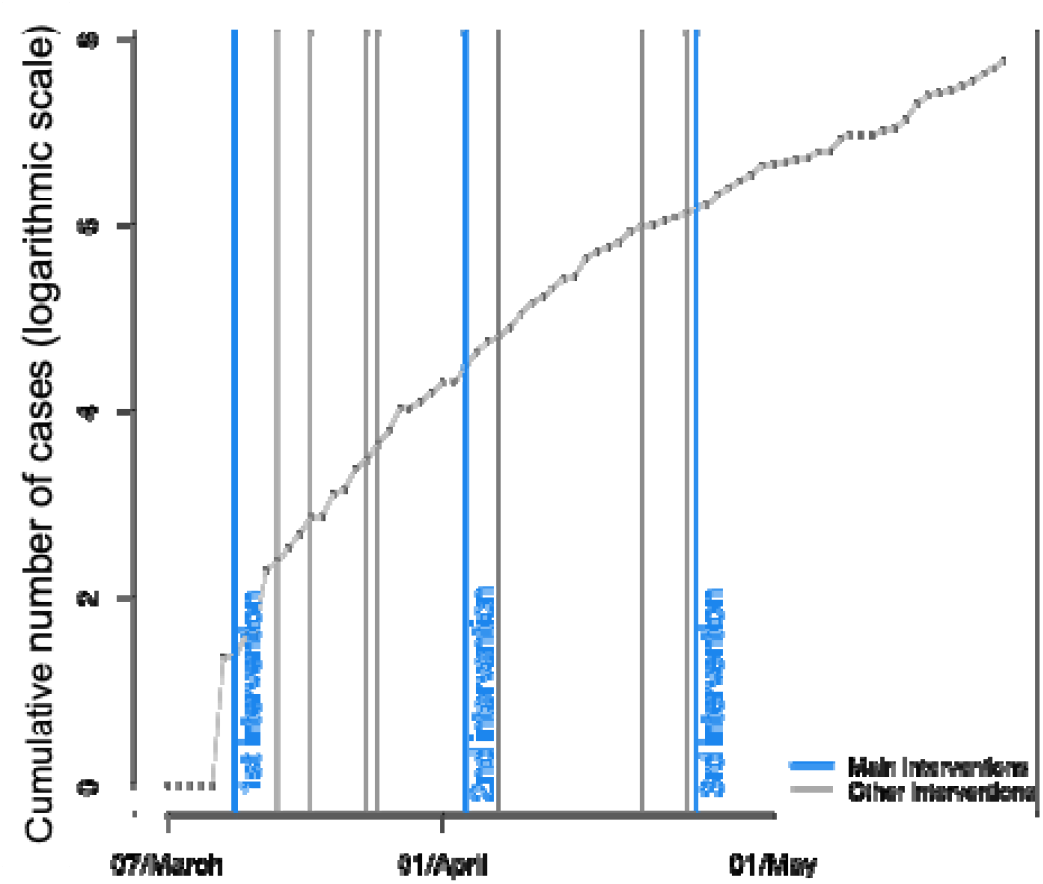
The cumulative cases of COVID-19 in the state of Goiás (logarithmic scale) and the 14 interventions made by the state’s government (until May 22). We chose the three main decrees capable of influencing public behavior as our three main interventions (blue lines). The first intervention was on March 13; The second intervention was on April 3; and the third intervention was on April 24. Other interventions are represented by the gray lines. Black dots represent confirmed cases.

The first intervention chosen was the first decree announced on March 13. In this decree the state declares a public health emergency and institutes strict social distancing measures, such as the shutdown of public and private events of any nature, including educational institutions at all levels, daycares, suspension of commercial activities such as malls, fairs, gyms, dental health services, religious meetings and all other non-essential services and activities *(31)*.

The second intervention chosen was the decree from April 3rd, which was already the eighth decree announced and the fourth relaxing the measures stated in the first one. This decree accumulates all prior flexibilizations, including reopening of religious activities, beauty salons, vegetable and fruit fairs, car workshops and restaurants on highways, administrative activities in public and private educational institutions *(31)*.

On April 19, the government launched a new decree extending the health emergency in Goiás for another 150 days. However, on April 24, they announced another decree altering the decree from 04/19, legislating on the private sphere as well (suspending activities of common use in closed condominiums), regulating a channel for reporting disobediences to any of the decrees, and legislating specific days for religious celebrations *(31)*. We chose the intervention on April 24 as the third changing point, as we see it to be more rigorous than previous ones.

### Simple SIR model: stationary spreading rate

We considered the initial onset transmission phase as being between March 6 and 20, approximately seven days before and after the first confirmed COVID-19 case in Goiás. Central epidemiological parameters for this model are the spreading rate (λ), recovery rate (μ), reporting delay *(D)* and number of initially infected people *(I*_0_) *(23)*. We chose informative log-normal priors of λ = 0.11 and μ = 0.11 (Table 1), as these priors cannot be estimated independently and these values maintain the effective growth rate (λ^*^ λ − μ) with a median of 0.19 and the basic reproduction number (R_0_ λ / μ) with a median of 2.72, consistent with global *(32)* and local *(33)* estimates. We chose for the reporting delay a prior that incorporates the virus’ incubation period between 1–14 days and the delay of infected people awaiting tests confirmation or medical appointments. Flat priors were chosen for the I_0_ and scale factor.

**Table 1.**
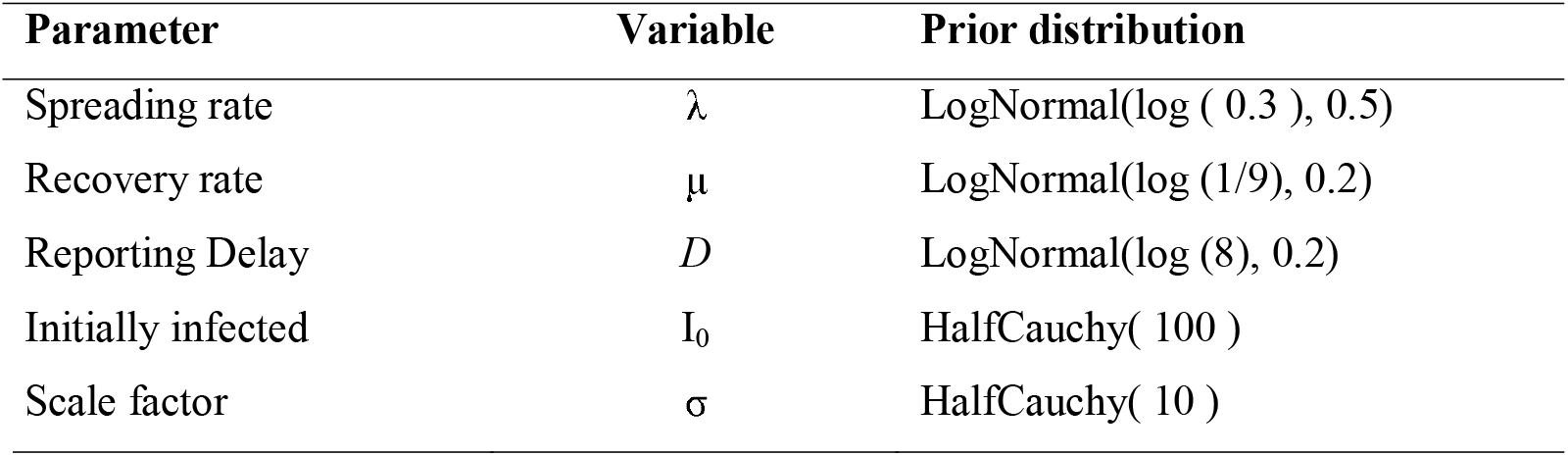
Priors for the simple SIR model with stationary spreading rate.

### Full SIR model: weekly reporting modulation and change points in spreading rate

To simulate the effect of governmental interventions, we use a SIR model with incorporated change points capable of altering the transmission rate *(23)*. The aim of interventions is to reduce the effective growth rate, thus if the rate becomes negative, new infections will begin to decrease. Dehning’s model assumes new spreading rates for each change point, inferred after supposed behavioral changes in the population.

We chose the same log-normal distributed priors for λ_0_, and *D* as in the simple model, with added parameters for the change points and their spreading rates (Table 2). We assumed the first government intervention reduced the spreading rate by 50% from the initial estimate λ_0_ = 0.3, so the prior for the first change point is λ_1_ ∼ logNormal (log (0.15), 0.5). Given the flexibilizations in the following decrees, we assumed the spreading rate would increase again by 15%, thus λ_2_ ∼ logNormal (log (0.22), 0.5). For the third intervention, there was more rigidity in the social distancing measures, which we presumed was embraced by the population. Therefore, we assumed the prior decreases the spreading rate and is closer (but slightly inferior) to the rate of the first intervention λ_3_ ∼ logNormal (log (0.11), 0.5).

**Table 2.** Priors for the full SIR model with change points and weekly reporting modulation.

| Parameter | Variable | Prior distribution |
| --- | --- | --- |
| Change points | $t_1$ | Normal(2020 / 03 / 06, 3) |
| | $t_2$ | Normal(2020 / 04 / 03, 1) |
| | $t_3$ | Normal(2020 / 04 / 24, 1) |
| Change duration | $\Delta t_i$ | LogNormal(log ( 3 ), 0.3) |
| Spreading rates | $\lambda_0$ | LogNormal(log ( 0.3 ), 0.5) |
| | $\lambda_1$ | LogNormal(log(0.22 ), 0.5) |
| | $\lambda_2$ | LogNormal(log( 0.22 ), 0.5) |
| | $\lambda_3$ | LogNormal(log (0.11), 0.5) |
| Recovery rate | $\mu$ | LogNormal(log (1/9), 0.2) |
| Reporting delay | $D$ | LogNormal(log (8), 0.2) |
| Weekly modulation amplitude | $f_w$ | Beta(mean = 0.7, std = 0.17) |
| Weekly modulation phase | $\phi_w$ | vonMises(mean = 0, $k = 0.01$) |
| Initially infected | $I_0$ | HalfCauchy( 100 ) |
| Scale factor | $\sigma$ | HalfCauchy( 10 ) |

We chose normal distributed priors for the timing of change points (Table 2). Respectively t_1_ ∼ Normal (2020/03/13, 3), t_2_ ∼ Normal (2020/04/03, 1) and t_3_ ∼ Normal (2020/04/24, 1), where 3, 1 and 1 are the respective transient days. Following the logic of the aforementioned decrees, we assumed the first intervention as a strict contact ban, the second as a mild contact ban and the third as again a strict contact ban. The change points take effect after a period of time (Δt_i_), for which we chose a median of 3 days. During these 3 days, spreading rates are expected to change for interventions to take effect. Furthermore, time is needed to ensure a smooth transition capable of absorbing the changes in the population’s behavior *(23)*.

Priors chosen for the recovery rate, reporting delay and initial number of infected people were the same as those applied in the simple SIR model (Table 1). The number of tests and reported cases varies throughout the week, with the number of records expected to be lower on weekends *(23)*. To implement the weekend effect in the model, we modulate the number of cases inferred by the absolute value of a sine function with the total period of 7 days *(23)*. This function was chosen by Dehning et al. (2020) because it is a non-symmetrical oscillation, adjusting the weekly variation of cases. We chose flat priors for the I_0_, scale factor and weekly modulation phase.

### Model comparison

Following *(23)*, we ran a model comparison using the leave-one-out (LOO) cross-validation method to avoid an over-fitting forecast. We compared four full SIR models with zero, one, two, and three change points, respectively. The full SIR model with three change points presented a better match between model and data (Table 3), as indicated by a lower LOO score. Full SIR models with zero, one and two change points performed poorly and will not be further discussed (but see Fig.S1-S2).

**Table 3.**
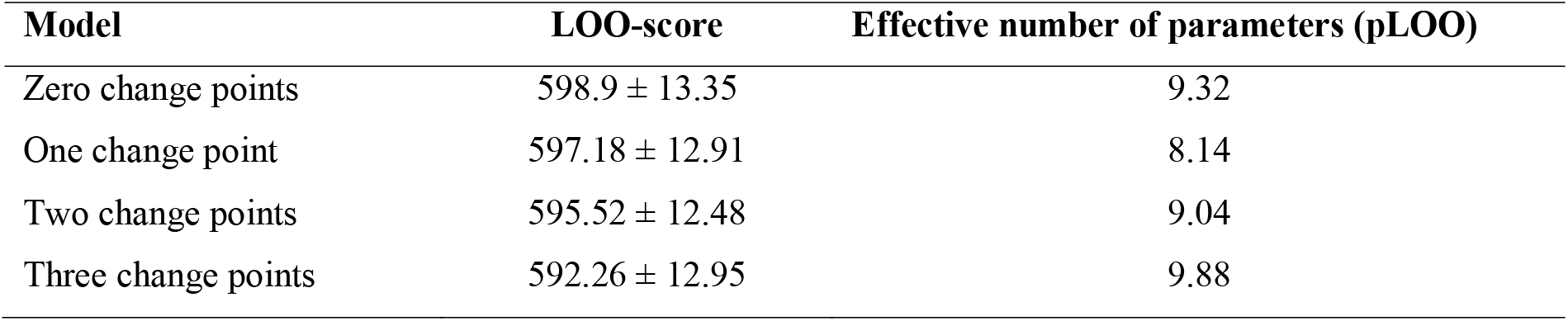
Leave-one-out (LOO) cross-validation for full SIR models (with weekend correction) with a different number of change points. Lower LOO-score indicates a better match between model and data.

| Model | LOO-score | Effective number of parameters (pLOO) |
| --- | --- | --- |
| Zero change points | $598.9 \pm 13.35$ | 9.32 |
| One change point | $597.18 \pm 12.91$ | 8.14 |
| Two change points | $595.52 \pm 12.48$ | 9.04 |
| Three change points | $592.26 \pm 12.95$ | 9.88 |

We also ran a model comparison with three change points but no weekend modulation and three sensitivity analyses by choosing wider priors to different parameters *(23)*. The model without a weekend modulation removes the assumption that daily reporting of new cases happens mainly during weekdays. The inferred parameters for this model are similar to the model that has the modulation, except for the number of initial infections (Fig. S3), but had a higher LOO-score when compared. For the sensitivity analysis, all parameters and priors were maintained exactly as the full SIR model, except were indicated. We ran a model with a prior four times wider for the reporting delay (Fig. S4), a model with a prior 14 days wide for the change times (Fig. S5) and a model with a prior four times wider for the change duration (Fig. S6). The full SIR model with three change points and weekly modulation again performed better than other models given the lower LOO-score (Table S1).

## Results

The daily reported cases in Goiás did not present an exponential curve in the simple SIR model with stationary spreading rate (Fig.2A), and the total reported cases (accumulated cases) show a tendency to be exponential (Fig.2B). The spreading rate was adjusted by the model as λ = 0.16 (95% credible interval (CI [0.07, 0.33];Fig. 2E)) and the effective growth rate as λ^*^ = 0.04 (Fig.2H), values lower than our prior. Further, μ and *D* histograms match the priors (gray line), as expected by the model *(23)*. The data for the initial phase is scarce and noisy, partly because the initial cases were not local infections, but of contaminated people arriving in Goiás. We expect the initial onset phase to be better explained by another phenomenon, such as migration of people coming from other states, not captured by our model. Thus, we will not be discussing these results further.

**Figure 2.**
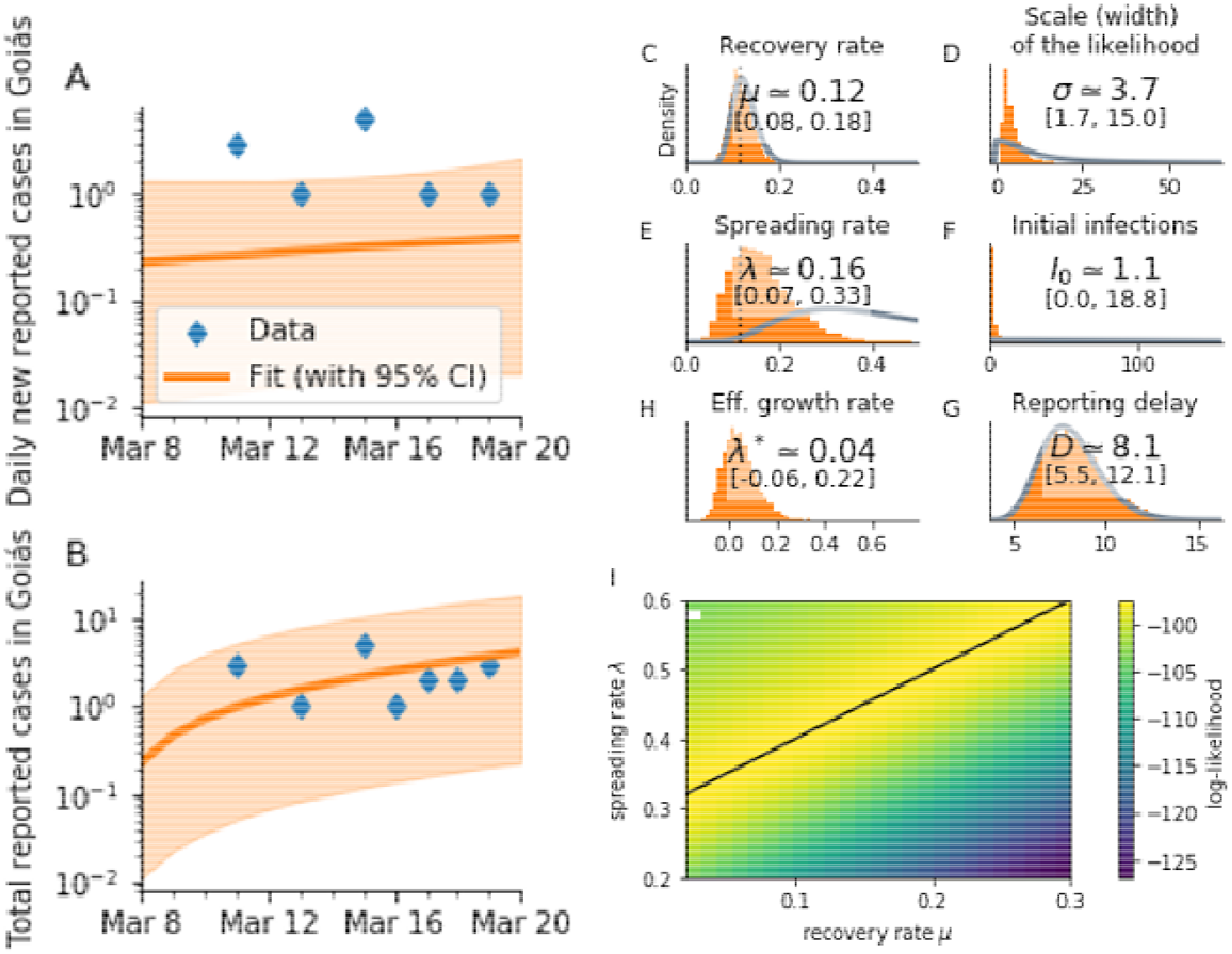
Results for the simple SIR model with stationary spreading rate during the initial onset period, March 8–20. A: Daily new reported cases in Goiás and; B: Total (cumulative) reported cases in Goiás. C-H: Inference of central epidemiological parameters: prior (gray) and posterior distributions (orange); C: Recovery rate µ; D: Scale-factor of the width of the likelihood distribution σ; E: estimated spreading rate λ; G: reporting delay *D*; I: Log-likelihood distribution for different combinations of λ and μ, the black line indicates a linear combination that yields the same maximal likelihood and the white dot indicates where inference did not converge.

In the full SIR model with change points and weekly reporting modulation, we found evidence of the influence of the three change points (Fig.3). First, the spreading rate decreased from λ_0_ = 0.28 (CI [0.12, 0.42]) to λ_1_ = 0.21 (CI [0.16, 0.29]). The date for the first change point was inferred as March 13 (CI [9, 21]), a date that marks the first state decree with strict contact ban measures. After this intervention, the effective growth rate was a median of λ_0_ – μ = 0.18 to median λ_1_-μ = 0.11, given μ was inferred as 0.10 (CI [0.07,0.14]).

**Figure 3.**
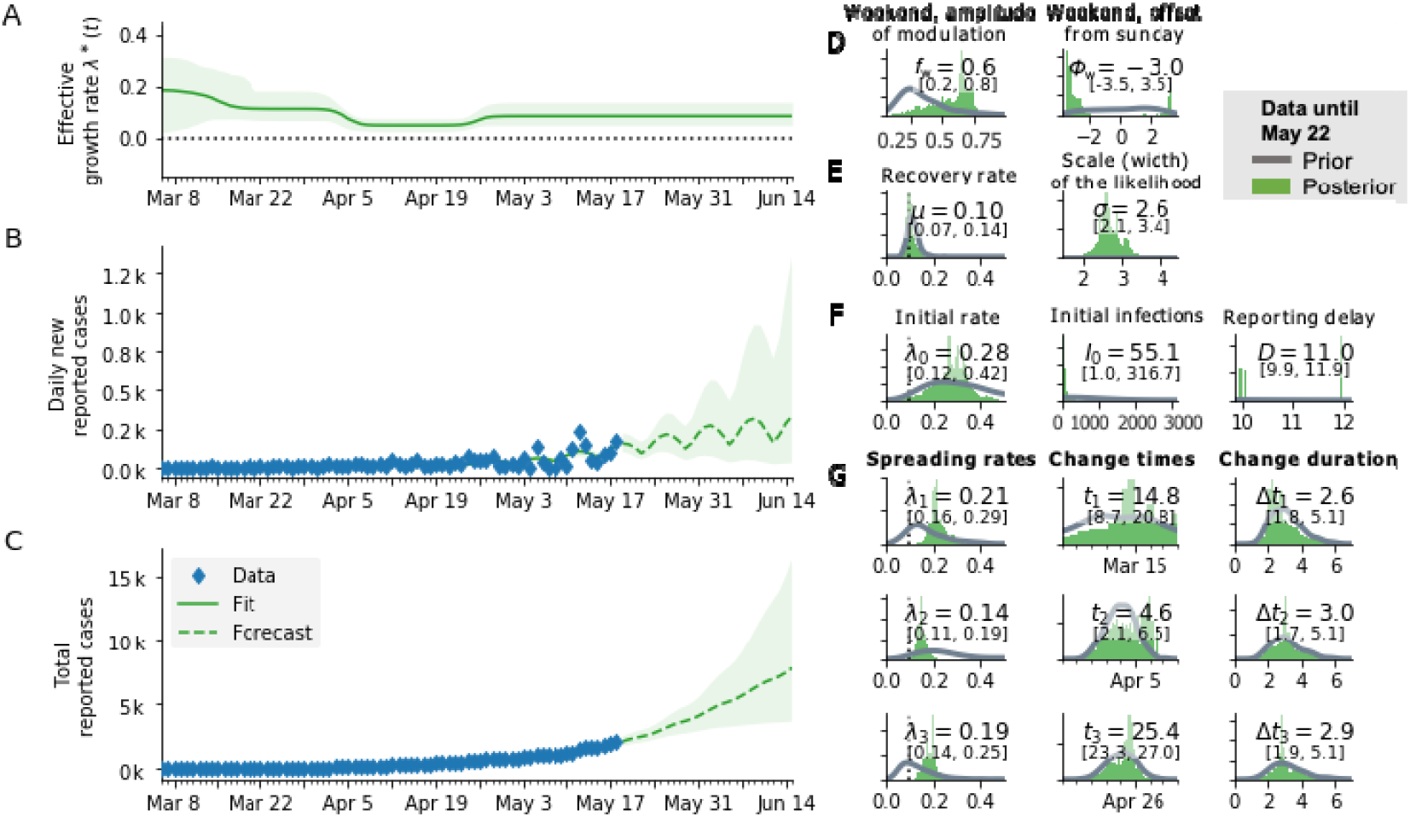
Results for the full SIR model with three change points and weekly reporting modulation. A: Estimate of the effective spreading rate; B: Daily new reported cases (blue diamonds) and the model(green solid line for median fit with 95% credible intervals). Green dashed line is the median forecast with 95% CI. C: Total reported cases and the model (color representation same as in B). D-F: Inference of central epidemiological parameters: prior (gray) and posterior distributions (green), inset values indicate the median and 95% CI of posteriors. G: Spreading rates, change times and change duration for the three change points, respectively.

At the second change point, λ_t_ decreased from λ_1_ = 0.21 to λ_2_ = 0.14 (CI [0.11, 0.19]), lower than assumed by our prior. This date was inferred as April 3 (CI [2, 6]), which marks the accumulation of flexibilizations from four decrees to first decree of March 13, including the reopening of religious events and fruit and vegetable fairs. After the second intervention, the median growth rate was λ_2_ – μ = 0.04, in the vicinity of a critical point (close to zero), but still positive.

The third change point increased λ_2_ = 0.14 to λ_3_ = 0.19 (CI [0.14, 0.25]). This change point was inferred to be April 24 (CI [23, 27]), a stricter decree compared to the previous ones. After this measure, the effective growth rate was of λ_3_ – μ = 0.09, indicating an increase in the growth, remaining above zero and thus not decreasing the number of new infections.

## Discussion

Given our results, the first two state-level interventions drastically reduced the COVID-19 spreading rate in Goiás. We expected the second intervention to increase the spreading rate, given prior relaxations, but the rate dropped. A plausible explanation could be that despite the relaxations of non-essential stores and services, the population obeyed social distancing. Nonetheless, the third intervention, although stricter than the second, brought the transmission rate back to a rate similar to that of the first intervention. We expected the third intervention to result in a similar transmission rate to that of the first intervention, but in the model, the transmission rate increased. This result probably reflects an accumulation of all fourteen state-level decrees and the population’s fatigue of being isolated, which probably resulted in more people eventually circulating in public spaces.

The transmission rates found in our model match the patterns found in a study that calculated time-series of the effective reproductive number *(Rt)* in Goiás (Diniz-Filho, Jardim, Toscano, & Rangel, 2020 [in press]). They found *Rt* to be around 2.0 in mid-March, dropping to approximately 1.2 in mid-April and increasing slowly to 1.4-1.5 in May. If we convert our transmission rates to R *(R = 1 +* λ *\* 5.2*; where 5.2 is the serial interval) we get R = 2.1 for the first intervention in March 13, R = 1.2 for the second intervention in April 3 and R = 1.5 for the third intervention in April 24. Both our models reflect the effects of the early social distancing measures implemented in Goiás in reducing the diseases’ onset transmission.

When this model was applied to Germany it demonstrated that following a gradual linear path of interventions, first banning major public events, later announcing mild social distancing measures and finally a strict contact ban *(23)*, aided in decreasing the transmission rate, bringing it to almost zero. Goiás followed an almost inversed path, imposing a first decree with strict social distancing measures at an early stage, but eventually reopening many specific services and activities. Nonetheless, if no social distancing measure had been imposed in Goiás, up to 62% of the population would have been infected by June 2^nd^ *(35)*, representing approximately 4 million people in the state. Further, it was also estimated that the interventions in Goiás prevented between 2.834 and 3.407 COVID-19 deaths *(35)*.

Although state-level interventions succeeded in decreasing the transmission rate, it remained high and exponential. Thus, other stricter interventions were made necessary to avoid the growth of new cases and a collapse in the health system. Nonetheless, more restrictive measures for the containment of COVID-19 were not adopted and were only discussed again in late June, when confirmed cases spiked. Our model forecasted for June 14 approximately 8.187 total reported cases (Fig. 3C), at that date, the state registered 7.944 confirmed cases *(30)*, a difference of 243 cases that could be explained by under testing and reporting delays.

Because no countries reached herd immunity *(8)*, second waves of infections are expected as well as more interventions to control them. Unfortunately, the accordion effect in COVID-19 spreading rate is not exclusive to Goiás. There have been new local outbreaks in 11 European countries, regions that previously lowered infection rates and were starting to lift restrictions *(36)*. Melbourne, in Australia, had to reimpose stay-at-home measures after a high increase of positive cases, including a border closure *(37)*. Governmental interventions need to be taken seriously by the population in order for them have the proposed outcome. Our results reflect the population’s disregard with the measures imposed. In contrast, countries with non-compulsory measures, such as Japan and Uruguay, experienced relatively low numbers of confirmed cases and deaths, as population self-isolated *(38, 39)*.

The COVID-19 outbreak poses itself as the biggest public health challenge in the last 100 years. Many countries around the world took drastic measures of social distancing and even complete lockdowns to contain it. Biomedical research on COVID-19 has boosted in the last six months and had many advances in clinical testing, drug repurposing and candidate vaccines *(40)*. While no effective treatment is made available, the better and safest way to successfully fight this pandemic is still social distancing and isolation, perhaps for an undetermined period of time. Governments (on any level) need cooperation from its citizens to succeed in containing COVID-19.

## Data Availability

All data used in this work is publicly available by the Goias State Health Department.

http://covid19.saude.go.gov.br/

## Authors’ contributions

All authors conceived and designed the study. TFR curated the data. CMB performed the analysis and wrote the first draft. All authors provided critical feedback, revised and approved the manuscript’s final version.

## Acknowledgments

We thank Mario Joaquim dos Santos Neto for the discussions and comments on the Brazilian legal system. We thank the Goiás State and Goiânia City Health Departments for support and access to original data.

